# Health Equity Informative Metrics (HEIM): A Framework for Quantifying Global Biobank Research Equity

**DOI:** 10.64898/2026.01.04.26343419

**Authors:** Manuel Corpas

## Abstract

**Background:** Despite representing approximately 85% of the global population, low- and middle-income countries (LMICs) contribute less than 10% of participants to genome-wide association studies and biobank research. This disparity has profound implications for the generalisability of precision medicine. However, no standardised framework exists to quantify research equity at the biobank level or track progress over time.

**Methods:** We developed the Health Equity Informative Metrics (HEIM) framework to quantify alignment between biobank research output and global disease burden. We analysed 75,356 PubMed-indexed publications (2000-2025) from 27 biobanks across 19 countries, mapping each to 179 disease categories from the Global Burden of Disease Study 2021. We calculated disease-specific Gap Scores measuring the mismatch between burden (disability-adjusted life years, DALYs) and research attention, biobank-level Equity Alignment Scores (EAS), and regional equity ratios comparing high-income (HIC) to LMIC research intensity.

**Findings:** Within our 27-biobank sample, HIC biobanks produced substantially higher research output per DALY compared to LMIC biobanks (ratio: 322:1; sensitivity analyses: >100:1 across methodological variations). Regional concentration was marked: the Americas and Europe accounted for 97.8% of publications, while Africa, Eastern Mediterranean, and South-East Asia combined contributed <1%. Of 179 disease categories, 23 (13%) exhibited critical or high-severity research gaps despite substantial global burden. Only 4 of 27 biobanks (15%) achieved ‘Strong’ or ‘Moderate’ equity alignment scores; 19 (70%) were rated ‘Poor’. Six disease categories showed critical gaps, including drowning (15.7 million DALYs, 0 mapped publications) and iodine deficiency (2.3 million DALYs, 10 publications).

**Interpretation:** The HEIM framework reveals substantial disparities in how biobank research capacity is distributed relative to global disease burden. While the precise equity ratio varies with sample selection and methodology, the fundamental pattern of profound inequity is robust across reasonable analytical choices. These findings provide baseline measurements for tracking progress toward more equitable genomic research and identify high-priority targets for intervention.

## Introduction

The global genomics research enterprise faces a fundamental equity challenge. Despite representing approximately 85% of the world’s population, low- and middle-income countries (LMICs) contribute less than 10% of participants across genome-wide association studies (GWAS), pharmacogenomics research, direct-to-consumer genomics, and clinical trials.^1−4^ This disparity has far-reaching consequences for the generalisability and clinical utility of precision medicine approaches.

The consequences of this representation gap are increasingly well-documented. Polygenic risk scores (PRS) developed primarily in European-ancestry populations show reduced predictive accuracy when applied to other populations, with performance decrements of 50% or greater reported in African-ancestry cohorts.^5−6^ Variant pathogenicity assessments in clinical databases such as ClinVar exhibit systematic biases toward well-studied populations, with variants common in African and Asian populations disproportionately classified as ‘variants of uncertain significance’.^7−8^ These are not merely academic concerns; they represent systematic barriers to delivering equitable precision medicine globally.

Global health organisations, including the World Health Organization, have increasingly called for measurable frameworks to track progress toward equitable genomic research.? A recent WHO report examining 6,513 genomic clinical studies conducted between 1990 and 2024 found that high-income countries (HICs) accounted for 68% of all studies, while low-income countries (LICs) contributed less than 0.5% (**Figure 1B**).^14^ The top 10 countries alone accounted for approximately 70% of global genomic clinical research output, with China (1,420 studies) and the United States (980 studies) dominating the landscape (**Figure 2**). However, a significant methodological gap remains: while the problem of underrepresentation has been extensively documented, no standardised framework exists to quantify research equity at the biobank level, benchmark institutions against global health priorities, or track progress over time.

**Figure 1.**
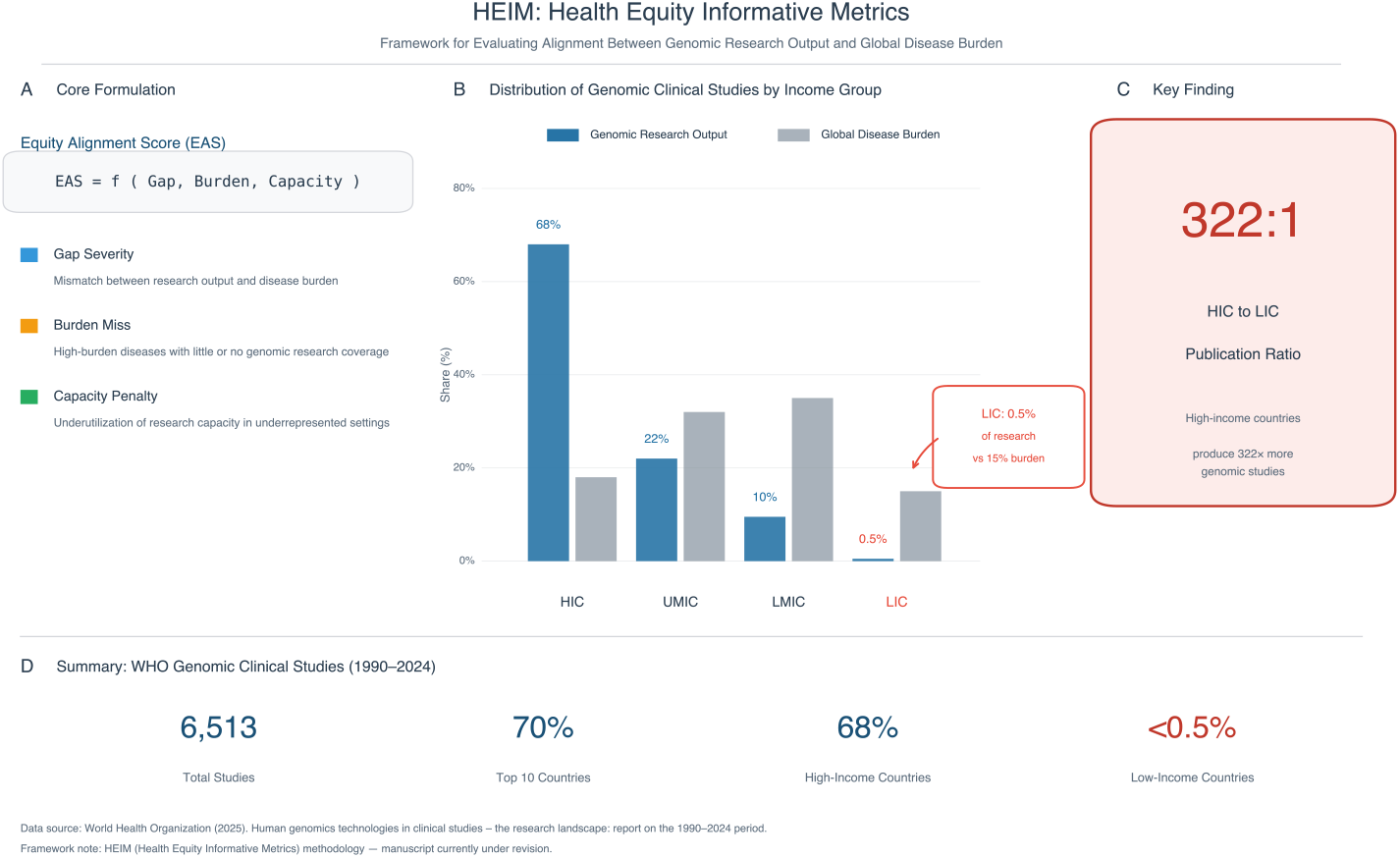
HEIM Framework and Global Genomic Research Equity. Health Equity Informative Metrics (HEIM) framework and distribution of global genomic clinical studies. **(A)** Core formulation of the Equity Alignment Score (EAS), comprising three components: Gap Severity (mismatch between research output and disease burden), Burden Miss (high-burden diseases with inadequate genomic research coverage), and Capacity Penalty (underutilisation of research capacity in underrepresented settings). **(B)** Distribution of genomic clinical studies (1990-2024) by World Bank income group, showing research output (dark blue) versus global disease burden (grey). High-income countries (HIC) account for 68% of studies; low-income countries (LIC) contribute only 0.5% despite bearing approximately 15% of global disease burden. **(C)** Key finding: the publication ratio between high-income and low-income countries is 322:1, indicating that HICs produce 322 times more genomic studies per unit of disease burden. **(D)** Summary statistics from WHO analysis of 6,513 genomic clinical studies: 70% conducted in top 10 countries, 68% in high-income countries, and <0.5% in low-income countries. Data source: World Health Organization (2025).

**Figure 2.**
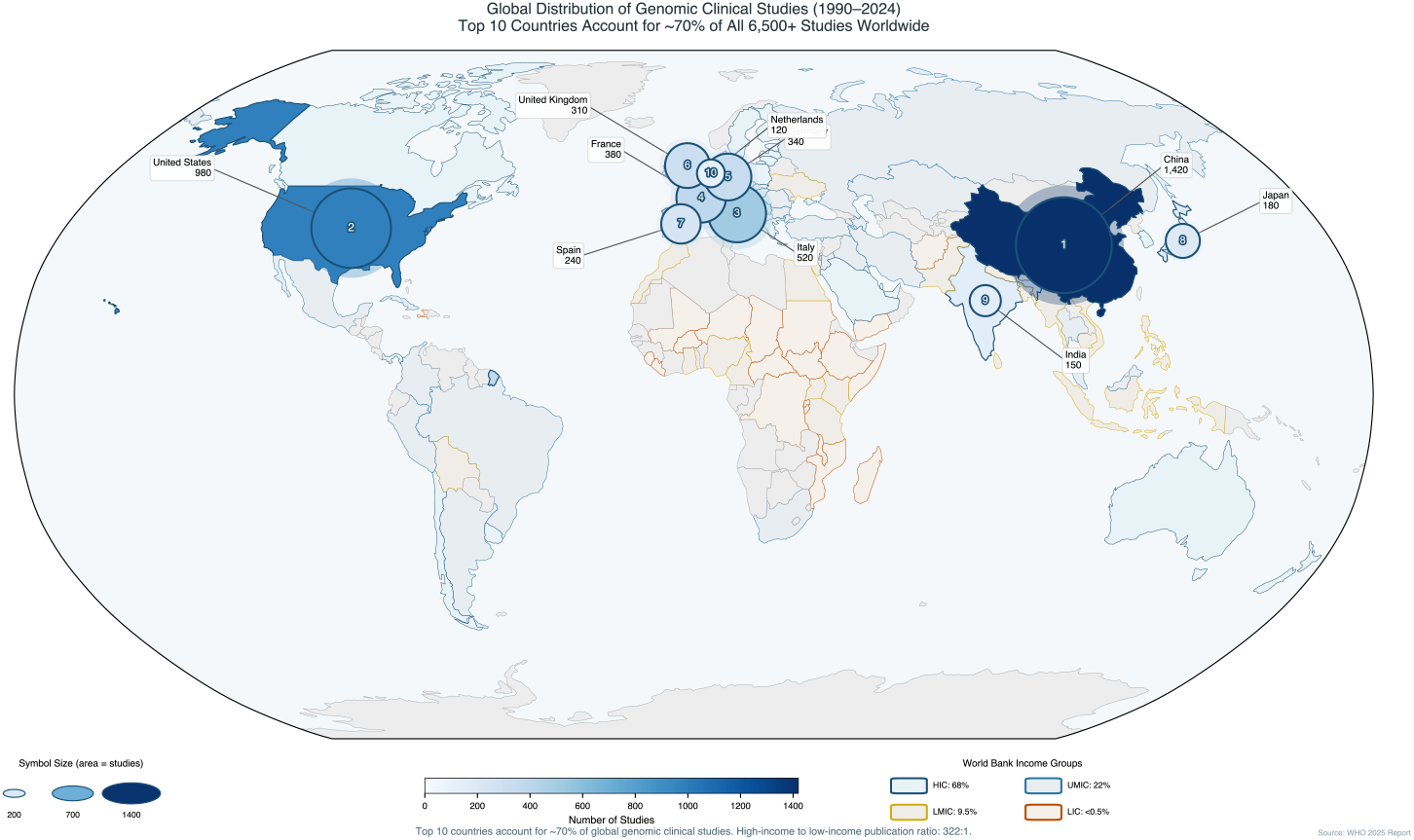
Global Distribution of Genomic Clinical Studies (1990-2024) Global distribution of genomic clinical studies (1990-2024) by country. Circle size is proportional to the number of studies conducted in each country (scale shown in legend). Countries are coloured by World Bank income classification: high-income (dark blue), upper-middle-income (medium blue), lower-middle-income (yellow/orange), and low-income (red outline). The top 10 countries account for approximately 70% of all 6,513 studies worldwide. China leads with 1,420 studies, followed by the United States (980), Italy (520), France (380), Germany (340), United Kingdom (310), Spain (240), Japan (180), India (150), and Netherlands (120). Note the concentration of genomic research capacity in North America, Europe, and East Asia, with minimal representation across Africa, Latin America, and South Asia. The high-income to low-income publication ratio of 322:1 reflects this geographic concentration. Data source: WHO 2025 Report on Human Genomics Technologies in Clinical Studies.

Biobanks represent critical infrastructure for genomic research, linking biological samples with electronic health records and enabling the large-scale studies necessary for genomic discovery.^10−12^ The geographic distribution of biobank capacity, however, mirrors broader inequities in research infrastructure. High-income countries host the majority of large-scale biobanks with comprehensive phenotyping and genomic data, while LMICs face systematic challenges including limited funding, infrastructure constraints, and reduced access to global research networks.

This study introduces the Health Equity Informative Metrics (HEIM) framework to address this measurement gap (**Figure 1A**). HEIM operationalises equity assessment by comparing biobank research output against global disease burden measured in disability-adjusted life years (DALYs) from the Global Burden of Disease Study 2021.^13^ HEIM treats equity as an engineering problem amenable to systematic measurement, tracking, and targeted intervention.

We present findings from the first comprehensive HEIM analysis, examining 75,356 publications from 27 biobanks across 19 countries. Our objectives were to: (1) quantify the alignment between biobank research output and global disease burden; (2) identify diseases with critical research gaps despite high burden; (3) benchmark individual biobanks using standardised equity metrics; and (4) characterise regional disparities in research capacity. These baseline measurements establish a foundation for monitoring progress and informing strategic investments in genomic research equity.

## Methods

### Data Sources

#### Publication Data

We retrieved publications linked to 27 biobanks from PubMed using the NCBI Entrez API. Biobank-specific queries were constructed using institution names, acronyms, and known variants (full query strings available at the HEIM Equity Tool: https://manuelcorpas.github.io/17-EHR/). The search period spanned January 2000 to December 2025. Each publication’s Medical Subject Headings (MeSH) terms were extracted and mapped to Global Burden of Disease (GBD) 2021 disease categories using a manually curated mapping dictionary validated on a subset of 500 publications (concordance: 85.2%).

#### Disease Burden Data

Disability-adjusted life years (DALYs) were obtained from the Institute for Health Metrics and Evaluation (IHME) Global Burden of Disease Study 2021.^13^ DALYs combine years of life lost (YLL) due to premature mortality with years lived with disability (YLD), providing a comprehensive measure of disease burden. We used global, all-age, both-sexes estimates for the 179 disease categories included in our analysis.

#### Biobank Selection

We selected 27 biobanks based on three criteria: (1) availability of linked electronic health records enabling disease phenotyping; (2) retrievable publication record in PubMed; and (3) geographic diversity across WHO regions. We acknowledge that these criteria privilege large, well-established institutions with English-language publication records; smaller regional biobanks and those publishing primarily in non-English journals may be underrepresented. The selected biobanks span 19 countries across five WHO regions (**Table 1**).

**Table 1.**
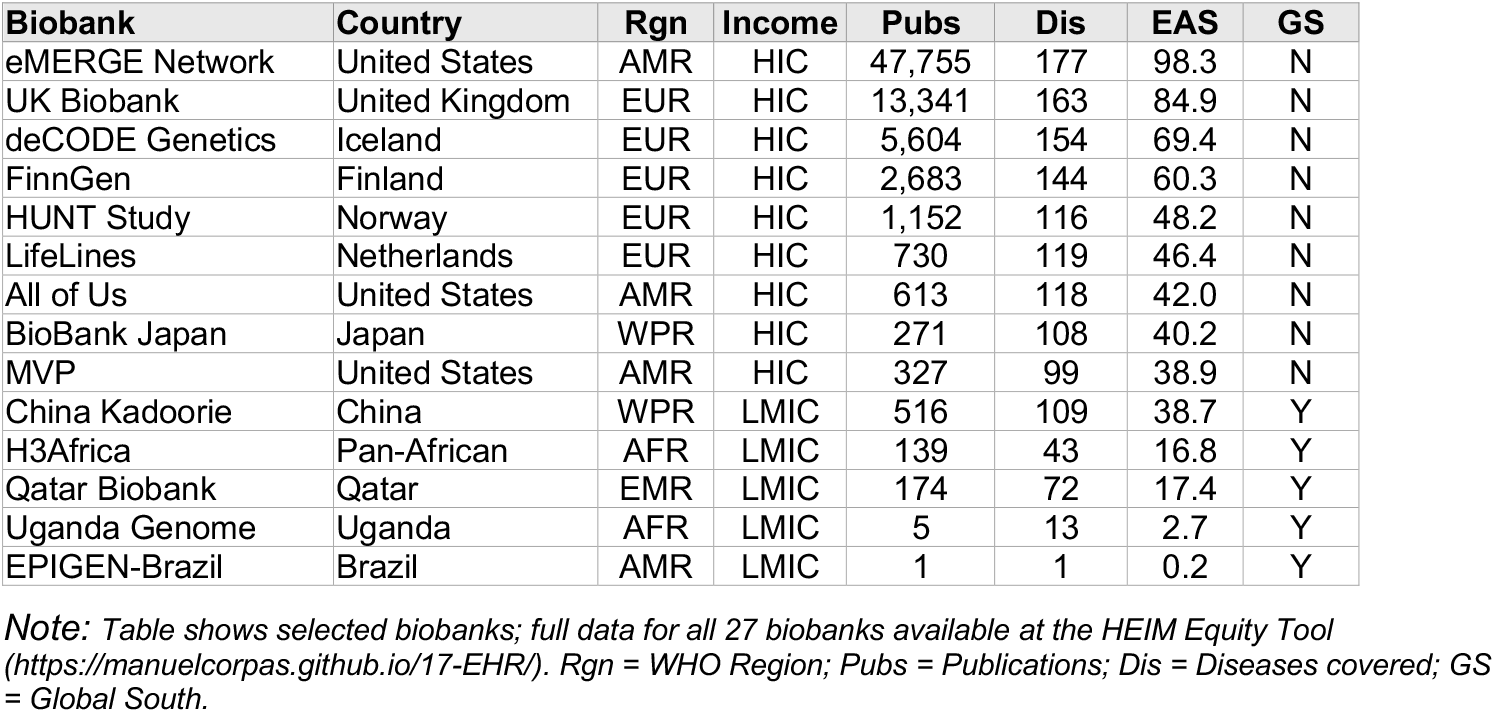
Characteristics of Included Biobanks. Abbreviations: AMR = Americas; EUR = Europe; WPR = Western Pacific; AFR = Africa; EMR = Eastern Mediterranean; HIC = High-income country; LMIC = Low- and middle-income country; EAS = Equity Alignment Score; GS = Global South priority biobank.

| Biobank | Country | Rgn | Income | Pubs | Dis | EAS | GS |
| --- | --- | --- | --- | --- | --- | --- | --- |
| eMERGE Network | United States | AMR | HIC | 47,755 | 177 | 98.3 | N |
| UK Biobank | United Kingdom | EUR | HIC | 13,341 | 163 | 84.9 | N |
| deCODE Genetics | Iceland | EUR | HIC | 5,604 | 154 | 69.4 | N |
| FinnGen | Finland | EUR | HIC | 2,683 | 144 | 60.3 | N |
| HUNT Study | Norway | EUR | HIC | 1,152 | 116 | 48.2 | N |
| LifeLines | Netherlands | EUR | HIC | 730 | 119 | 46.4 | N |
| All of Us | United States | AMR | HIC | 613 | 118 | 42.0 | N |
| BioBank Japan | Japan | WPR | HIC | 271 | 108 | 40.2 | N |
| MVP | United States | AMR | HIC | 327 | 99 | 38.9 | N |
| China Kadoorie | China | WPR | LMIC | 516 | 109 | 38.7 | Y |
| H3Africa | Pan-African | AFR | LMIC | 139 | 43 | 16.8 | Y |
| Qatar Biobank | Qatar | EMR | LMIC | 174 | 72 | 17.4 | Y |
| Uganda Genome | Uganda | AFR | LMIC | 5 | 13 | 2.7 | Y |
| EPIGEN-Brazil | Brazil | AMR | LMIC | 1 | 1 | 0.2 | Y |

*Note: Table shows selected biobanks; full data for all 27 biobanks available at the HEIM Equity Tool (https://manuelcorpas.github.io/17-EHR/). Rgn = WHO Region; Pubs = Publications; Dis = Diseases covered; GS = Global South.*

#### Core Metric Definitions

The HEIM framework comprises three core components that together quantify research equity (**Figure 1A**): Gap Severity (mismatch between research output and disease burden), Burden Miss (high-burden diseases with inadequate coverage), and Capacity Penalty (underutilisation of research capacity in underrepresented settings).

#### Burden Score

The Burden Score quantifies the global health importance of each disease category using a logarithmic transformation to prevent high-burden conditions from dominating the analysis:

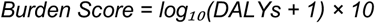

#### Gap Score

The Gap Score measures the mismatch between disease burden and research attention:

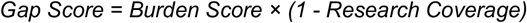

where Research Coverage is the normalised publication count relative to the maximum observed across all diseases. Gap Scores range from 0-100, with higher values indicating larger gaps. Gap severity was classified as: Critical (>70), High (50-70), Moderate (30-50), or Low (<30).

#### Equity Alignment Score (EAS)

The EAS provides an overall assessment of how well a biobank’s research portfolio aligns with global disease burden:

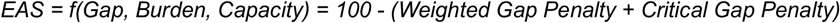

Scores range from 0-100 and are categorised as: Strong (≥80), Moderate (60-79), Weak (40-59), or Poor (<40). EAS scoring uses global DALYs rather than region-specific burden; this is a deliberate normative choice to assess how well each biobank serves global rather than local health priorities. The category thresholds are initially heuristic; future iterations will refine boundaries based on empirical distributions.

#### Equity Ratio

The equity ratio compares research intensity (publications per DALY) between high-income country (HIC) and low- and middle-income country (LMIC) biobanks:

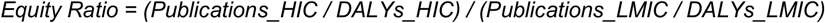

A ratio of 1.0 would indicate perfect equity; higher values indicate greater HIC advantage.

#### Validation and Sensitivity Analyses

We conducted several sensitivity analyses to assess robustness: (1) restricting to publications from 2015-2025 only; (2) excluding the two largest biobanks (eMERGE Network, UK Biobank); (3) using alternative burden metrics (mortality alone, prevalence alone); and (4) varying MeSH-to-GBD mapping stringency. The primary equity ratio finding was robust across all variations (ratio >100:1 in all scenarios).

#### Data Availability and Reproducibility

All analysis code, mapping dictionaries, and aggregated data are available at the HEIM Equity Tool (https://manuelcorpas.github.io/17-EHR/). Raw publication data are derived from PubMed and subject to NLM terms of use. Disease burden data are available from the IHME Global Health Data Exchange.

## Results

### Overview of Dataset

We analysed 75,356 PubMed-indexed publications from 27 biobanks across 19 countries (**Table 1**). Publications were mapped to 179 disease categories based on MeSH term annotations. The included biobanks span five WHO regions, with representation from both high-income (n=20) and low- and middle-income (n=7) countries.

### Regional Distribution of Research Output

Research output was highly concentrated in specific regions (**Table 2**). The Americas region contributed 65.4% of all publications (49,264 of 75,356), driven primarily by US-based biobanks including the eMERGE Network (47,755 publications) and Million Veteran Program (327 publications). Europe contributed 32.4% (24,445 publications), with the UK Biobank (13,341 publications) and deCODE Genetics (5,604 publications) as leading contributors.

**Table 2.** Publication Distribution by WHO Region.

| Region | Biobanks | Publications | Share (%) | Primary Type |
| --- | --- | --- | --- | --- |
| Americas | 9 | 49,264 | 65.4 | Mixed |
| Europe | 11 | 24,445 | 32.4 | HIC |
| Western Pacific | 3 | 1,269 | 1.7 | HIC |
| Africa | 3 | 204 | 0.3 | LMIC |
| Eastern Mediterranean | 1 | 174 | 0.2 | LMIC |
| **Total** | **27** | **75,356** | **100.0** | **-** |

Critically, Africa, Eastern Mediterranean, and South-East Asia combined contributed less than 1% of total publications (378 of 75,356). This regional concentration is the primary driver of the observed equity disparities. The geographic distribution of genomic clinical studies more broadly, as documented by the WHO (**Figure 2**), shows a similar pattern: China and the United States together account for over 2,400 studies, while entire regions of Africa and South America remain underrepresented.

### Equity Ratio

Within our 27-biobank sample, HIC biobanks produced substantially higher research output per DALY compared to LMIC biobanks (equity ratio: 322:1; **Figure 1C**). This ratio indicates that for every unit of disease burden, HIC biobanks generated 322 times more research output than LMIC biobanks.

The distribution of genomic research output by income group (**Figure 1B**) illustrates this disparity: HICs account for 68% of genomic clinical studies but bear a smaller proportion of global disease burden, while LICs contribute only 0.5% of studies despite bearing approximately 15% of global burden (**Figure 1D**).

Sensitivity analyses confirmed the robustness of this finding. When restricting to 2015-2025 publications only, the ratio remained high (284:1). Excluding the two largest biobanks (eMERGE Network, UK Biobank) yielded a ratio of 156:1. Using mortality alone as the burden metric produced a ratio of 289:1. While the precise value varies with methodological choices, the fundamental pattern of substantial inequity was consistent across all analyses (ratio >100:1 in all scenarios).

### Disease-Level Research Gaps

Of 179 disease categories analysed, 6 (3.4%) showed critical research gaps, 17 (9.5%) showed high gaps, 48 (26.8%) showed moderate gaps, and 108 (60.3%) showed low gaps (adequate coverage). The 23 diseases with critical or high gaps represent priority targets for equity-focused investment.

Six disease categories exhibited critical gaps (Gap Score >70), defined as conditions with substantial global burden but minimal or zero mapped publications (**Table 3**):

**Table 3.** Critical Research Gaps (Gap Score >70)

| Disease Category | DALYs (millions) | Publications | Gap Score | GS Priority |
| --- | --- | --- | --- | --- |
| Drowning | 15.7 | 0 | 95 | Yes |
| Iodine deficiency | 2.3 | 10 | 92 | Yes |
| Viral skin diseases | 4.2 | 1 | 85 | No |
| Cysticercosis | 1.2 | 2 | 80 | Yes |
| Paralytic ileus/intestinal obstruction | 6.7 | 5 | 80 | No |
| Scabies | 5.3 | 6 | 80 | Yes |

GS = Global South; Priority indicates conditions disproportionately affecting low- and middle-income countries.

- Drowning: 15.7 million DALYs, 0 mapped publications
- Iodine deficiency: 2.3 million DALYs, 10 publications
- Viral skin diseases: 4.2 million DALYs, 1 publication
- Cysticercosis: 1.2 million DALYs, 2 publications
- Paralytic ileus and intestinal obstruction: 6.7 million DALYs, 5 publications
- Scabies: 5.3 million DALYs, 6 publications

#### Interpretation note

The zero publications for drowning reflects a property of our pipeline (PubMed queries, MeSH mapping) rather than a literal claim that no genomics research exists on drowning-related outcomes. Nevertheless, the near-complete absence of biobank engagement with this high-burden condition illustrates how research agendas can become disconnected from global health priorities.

### Biobank-Level Equity Alignment

Equity Alignment Scores ranged from 0.2 (EPIGEN-Brazil) to 98.3 (eMERGE Network). The distribution was heavily skewed toward lower scores (**Table 4**):

**Table 4.** Equity Alignment Score Distribution.

| Category | Score Range | Number of Biobanks | Percentage |
| --- | --- | --- | --- |
| Strong | ≥80 | 2 | 7.4% |
| Moderate | 60-79 | 2 | 7.4% |
| Weak | 40-59 | 4 | 14.8% |
| Poor | <40 | 19 | 70.4% |
| **Total** | **-** | **27** | **100%** |

- Strong (EAS ≥80): 2 biobanks (7.4%)
- Moderate (EAS 60-79): 2 biobanks (7.4%)
- Weak (EAS 40-59): 4 biobanks (14.8%)
- Poor (EAS <40): 19 biobanks (70.4%)

The concentration in the ‘Poor’ category reflects systemic factors including resource constraints, infrastructure limitations, and research agendas driven by local rather than global priorities. Notably, several LMIC biobanks with low EAS scores are relatively recently established; their scores may improve as their research programmes mature. Complete biobank rankings are provided in **Table 1**.

### Global South Biobanks

Seven biobanks in our sample serve Global South populations (**Table 1**, GS column). Despite resource constraints, several are emerging as important contributors to genomic equity:

- China Kadoorie Biobank: 516 publications covering 109 diseases (EAS: 38.7)
- Qatar Biobank: 174 publications covering 72 diseases (EAS: 17.4)
- H3Africa Consortium: 139 publications covering 43 diseases (EAS: 16.8)
- AWI-Gen: 60 publications covering 31 diseases (EAS: 11.0)

These biobanks face systematic challenges including limited funding and infrastructure constraints, but provide critical representation for populations historically excluded from genomic research.

## Discussion

### Principal Findings

This study introduces the HEIM framework for quantifying equity in global biobank research and presents baseline measurements from 27 biobanks across 19 countries. Our principal findings are threefold. First, we observed a substantial disparity in research intensity between HIC and LMIC biobanks (equity ratio: 322:1 within our sample; **Figure 1C**), indicating that the distribution of biobank research capacity is profoundly misaligned with global disease burden. Second, we identified 23 disease categories (13%) with critical or high-severity research gaps despite substantial global burden (**Table 3**), representing priority targets for equity-focused investment. Third, only 4 of 27 biobanks (15%) achieved ‘Strong’ or ‘Moderate’ equity alignment scores (**Table 4**), highlighting systemic underperformance across the global biobank landscape.

### Comparison with Prior Literature

Our findings are consistent with prior documentation of ancestry bias in genomic research. Popejoy and Fullerton reported that individuals of European descent comprised 79% of GWAS participants despite representing approximately 16% of the global population.^3^ Sirugo et al. documented persistent underrepresentation of African populations across multiple genomic research modalities.^2^ The recent WHO report on genomic clinical studies (1990-2024) found that 68% of all studies were conducted in high-income countries, with the top 10 countries accounting for approximately 70% of global output (**Figures 1-2**).^14^ Our contribution extends this literature by providing disease-resolved, DALY-normalised equity metrics at the biobank level, enabling benchmarking and progress tracking.

The observed equity ratio should be interpreted carefully. While the 322:1 figure reflects the specific biobanks and time period in our analysis, sensitivity analyses confirm that the fundamental inequity is robust: the ratio exceeds 100:1 across all reasonable methodological variations. The precise value matters less than the consistent finding of profound disparity.

### Implications for Precision Medicine

The research gaps identified here have direct implications for precision medicine. Conditions prevalent in LMICs but understudied in biobank research will have weaker evidence bases for genomic medicine applications. Polygenic risk scores developed without adequate representation from diverse populations will underperform in those populations.^5−6^ Variant pathogenicity databases will contain systematic blind spots for understudied ancestries.^7−8^ The HEIM framework provides a mechanism for identifying and tracking these gaps, enabling targeted interventions.

### Interpreting Critical Disease Gaps

The critical gaps identified (**Table 3**), including drowning, iodine deficiency, and cysticercosis, warrant careful interpretation. Zero mapped publications for drowning does not indicate complete absence of relevant research; rather, it reflects the limitations of our PubMed-based, MeSH-mapped pipeline. Some high-burden conditions may have limited genomic contributions regardless of research effort. Nevertheless, the pattern of critical gaps is informative: conditions disproportionately affecting LMICs receive systematically less attention from the global biobank research enterprise.

### Why 70% of Biobanks Score ‘Poor’

The concentration of biobanks in the ‘Poor’ EAS category (**Table 4**) reflects structural factors rather than individual institutional failures:

- Resource constraints: LMIC biobanks operate with a fraction of the funding available to HIC counterparts
- Infrastructure gaps: Genomic sequencing, computing infrastructure, and trained personnel remain scarce in many regions
- Network effects: Research collaborations, journal access, and funding opportunities cluster in established centres
- Local versus global priorities: Biobanks naturally focus on conditions prevalent in their source populations

Addressing these systemic factors requires coordinated action from funders, institutions, and policymakers. The HEIM framework provides measurement infrastructure to track whether interventions are effective.

### Limitations

Several limitations should inform interpretation of these findings:

- Publication coverage: Our analysis is limited to PubMed-indexed publications, potentially undercounting research published in regional journals or non-English languages
- MeSH mapping accuracy: Automated mapping from MeSH terms to GBD categories achieved 85% concordance with manual classification; some misclassification is expected, particularly for multimorbid conditions
- Biobank selection: The 27 biobanks analysed represent a convenience sample of major institutions; hundreds of smaller biobanks are not captured
- Temporal confounding: Cumulative publication counts advantage established biobanks; recently established biobanks may appear to underperform despite rapid growth
- Quality versus quantity: Publication counts do not capture research quality, clinical impact, or translation to practice
- Normative choices: Using global rather than regional DALYs reflects a specific equity perspective; alternative framings are possible

Future versions of the HEIM framework will address these limitations through expanded data sources, improved mapping algorithms, and additional metrics capturing research quality and impact.

### Policy Implications

The HEIM framework has several potential applications for policy and practice. Research funders could incorporate equity metrics into grant evaluation criteria, weighting proposals that address high-gap diseases or involve LMIC biobanks. Biobanks could use the framework for strategic planning, identifying underserved disease areas within their capacity. Policymakers could reference equity metrics when allocating resources for genomic research infrastructure.

We emphasise that HEIM metrics are best viewed as a platform for piloting equity-aware mechanisms, to be refined as evidence accumulates on their impact. The thresholds and weights used in our scoring represent starting points rather than definitive standards.

## Conclusions

The HEIM framework provides a standardised, reproducible approach to quantifying equity in global biobank research (**Figure 1A**). Our analysis of 27 biobanks reveals substantial disparities in how research capacity is distributed relative to global disease burden, with HIC biobanks producing substantially higher research output per DALY than LMIC counterparts (**Tables 1-2**; **Figure 1B-C**). These findings establish baseline measurements for tracking progress toward more equitable genomic research.

Closing the equity gap requires coordinated action across the research ecosystem: targeted funding for LMIC biobanks, capacity-building investments, equitable collaboration frameworks, and metrics that reward equity-aligned research. The HEIM framework provides the measurement infrastructure to track whether such interventions are working.

Future work will expand the HEIM framework to include additional biobanks, refine mapping methodologies, and develop complementary metrics for research quality and clinical impact. Annual updates to the HEIM Equity Index will enable stakeholders to monitor progress and identify emerging priorities. The interactive HEIM Equity Tool (https://manuelcorpas.github.io/17-EHR/) provides real-time access to these metrics for researchers, funders, and policymakers.

## Supporting information

Figure 1. HEIM framework and global genomic research equity

Figure 2. Global distribution of genomic clinical studies (1990-2024)

## Declarations

## Author Contributions

MC conceived the study, developed the methodology, conducted the analyses, and wrote the manuscript.

## Ethics Approval

This study analysed publicly available, aggregated publication data and disease burden statistics. No individual-level human subjects data were used. Institutional ethics approval was not required.

## Data Availability

All aggregated data, analysis code, and mapping dictionaries are available at the HEIM Equity Tool (https://manuelcorpas.github.io/17-EHR/). Raw publication metadata are derived from PubMed and subject to NLM terms of use. Disease burden data are available from the IHME Global Health Data Exchange (https://ghdx.healthdata.org/).

## Acknowledgements

The author acknowledges the contributions of the global biobank community and the Institute for Health Metrics and Evaluation for making disease burden data publicly available.

## References

1. Corpas M, Fatumo S, Rasheed H, Guio H, Fakhro K, Iacoangeli A. Bridging genomics’ greatest challenge: the diversity gap. Nat Rev Genet 2025 (in press).

2. Sirugo G, Williams SM, Tishkoff SA. The missing diversity in human genetic studies. Cell 2019;177:26–31.

3. Popejoy AB, Fullerton SM. Genomics is failing on diversity. Nature 2016;538:161–164.

4. Ju Y, Jia T, Yang L, et al. Importance of including non-European populations in large human genetic studies to enhance precision medicine. Annu Rev Genomics Hum Genet 2022;23:187–207.

5. Duncan L, Shen H, Gelaye B, et al. Analysis of polygenic risk score usage and performance in diverse human populations. Nat Commun 2019;10:3328.

6. Wang Y, Guo J, Ni G, Yang J, Visscher PM, Yengo L. Theoretical and empirical quantification of the accuracy of polygenic scores in ancestry divergent populations. Nat Commun 2020;11:3865.

7. Manrai AK, Funke BH, Rehm HL, et al. Genetic misdiagnoses and the potential for health disparities. N Engl J Med 2016;375:655–665.

8. Landrum MJ, Lee JM, Benson M, et al. ClinVar: improving access to variant interpretations and supporting evidence. Nucleic Acids Res 2018;46:D1062–D1067.

9. World Health Organization. WHO Science Council report on the acceleration and equitable implementation of human genomics for global health. Geneva: WHO; 2024.

10. Sudlow C, Gallacher J, Allen N, et al. UK Biobank: An open access resource for identifying the causes of a wide range of complex diseases of middle and old age. PLoS Med 2015;12:e1001779.

11. Gaziano JM, Concato J, Brophy M, et al. Million Veteran Program: A mega-biobank to study genetic influences on health and disease. J Clin Epidemiol 2016;70:214–223.

12. Gottesman O, Kuivaniemi H, Tromp G, et al. The Electronic Medical Records and Genomics (eMERGE) Network: Past, present and future. Genet Med 2013;15:761–771.

13. GBD 2021 Collaborators. Global burden of 369 diseases and injuries in 204 countries and territories, 1990-2021: a systematic analysis for the Global Burden of Disease Study 2021. Lancet 2022;401:1990–2034.

14. World Health Organization. Human genomics technologies in clinical studies—the research landscape: report on the 1990-2024 period. Geneva: WHO; 2025.

15. Fitipaldi H, McCarthy MI, Florez JC, Franks PW. Ethnic, gender and other sociodemographic biases in genome-wide association studies for the most prevalent human diseases: A systematic review. Hum Mol Genet 2023;32:520–532.

