## Supplementary material for "Health Equity Informative Metrics (HEIM): A Framework for Quantifying Global Biobank Research Equity": Figure 1. HEIM framework and global genomic research equity

### HEIM: Health Equity Informative Metrics

Framework for Evaluating Alignment Between Genomic Research Output and Global Disease Burden

#### A Core Formulation

##### Equity Alignment Score (EAS)

```
EAS = f ( Gap, Burden, Capacity )
```

**Gap Severity**

Mismatch between research output and disease burden

**Burden Miss**

High-burden diseases with little or no genomic research coverage

**Capacity Penalty**

Underutilization of research capacity in underrepresented settings

#### B Distribution of Genomic Clinical Studies by Income Group

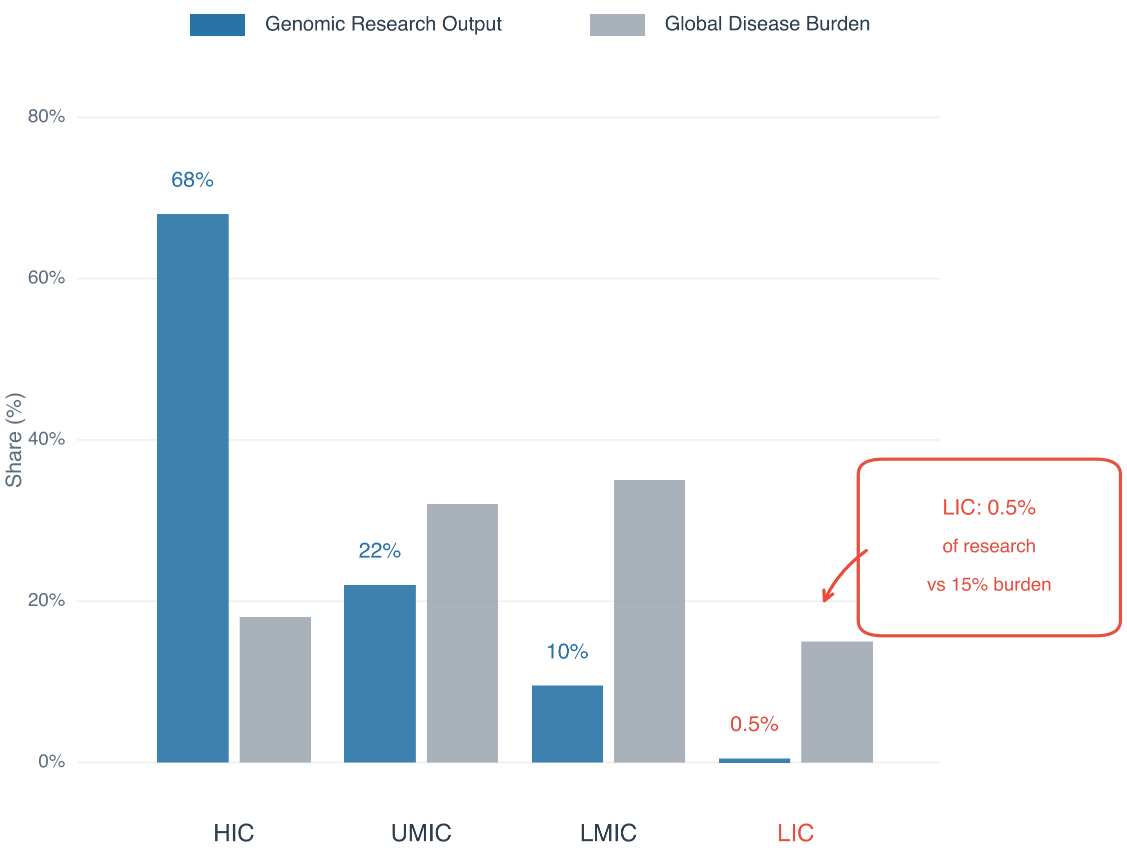


#### C Key Finding

**322:1**

HIC to LIC

Publication Ratio

High-income countries

produce 322× more genomic studies

#### D Summary: WHO Genomic Clinical Studies (1990–2024)

| 6,513 | 70% | 68% | <0.5% |
|---|---|---|---|
| Total Studies | Top 10 Countries | High-Income Countries | Low-Income Countries |

Data source: World Health Organization (2025). Human genomics technologies in clinical studies – the research landscape: report on the 1990–2024 period.

Framework note: HEIM (Health Equity Informative Metrics) methodology — manuscript currently under revision.
