## Supplementary material for "Health Equity Informative Metrics (HEIM): A Framework for Quantifying Global Biobank Research Equity": Figure 2. Global distribution of genomic clinical studies (1990-2024)

### Global Distribution of Genomic Clinical Studies (1990–2024)
### Top 10 Countries Account for ~70% of All 6,500+ Studies Worldwide

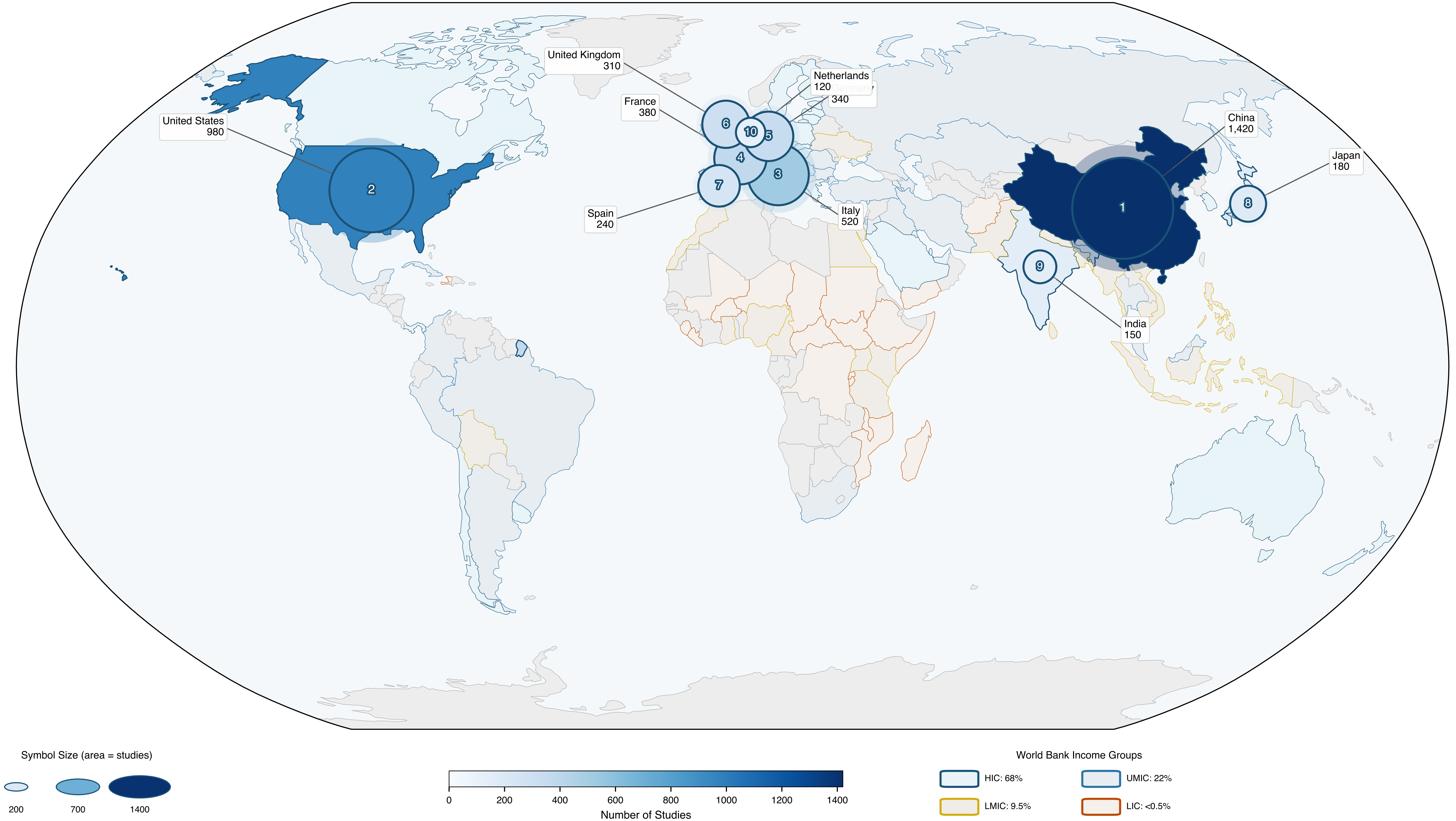


Top 10 countries account for ~70% of global genomic clinical studies. High-income to low-income publication ratio: 322:1.

Source: WHO 2025 Report
